# A Multi-Task Deep Learning Model for Pediatric Echocardiography Analysis

**DOI:** 10.1101/2025.10.27.25338912

**Authors:** Cho Joseph, Mathur Mrudang, Kaur Dhamanpreet, Duda Matthew, Dahlan Adil, Krishnan Aravind, Leipzig Matthew, Shad Rohan, Alexander K. Gonzalez, Logan Joseph, Seidman Christa, Fong Robyn, Kumar Abhinav, Zakka Cyril, Curtis P. Langlotz, Matthew A. Jolley, Hiesinger William

## Abstract

**Background:** Congenital heart defects afflict nearly 1% of all births worldwide. While deep learning algorithms have shown significant promise in automating and improving adult echocardiography analysis, similar progress has not been observed in pediatric echocardiography. Specifically, existing pediatric-based models are limited to single tasks and specific echocardiographic views. To address this, we introduce EchoAI-Peds, the first multi-task deep learning model for pediatric echocardiography. Our model was developed using the most comprehensive set of pediatric labels to date and is designed to integrate information from multiple echocardiographic views simultaneously.

**Methods:** A video-based vision transformer was trained to simultaneously detect 28 congenital heart defects, structural and functional abnormalities, repairs, and interventions directly from complete pediatric echocardiography studies with multiple videos. During inference, our model integrates information from all available views to produce unified study-level predictions. Our model was developed using over 700,000 videos derived from more than 11,000 studies at Stanford Medicine. Model efficacy was tested on an internal held-out dataset. In addition, model generalizability was tested on a spatially and temporally distinct patient cohort at the Children’s Hospital of Philadelphia.

**Results:** Our model achieved macro-averaged AUROC values of 0.91 (95% CI: 0.90-0.92) and 0.89 (95% CI: 0.88-0.90) on the internal and external test sets, respectively. Moreover, our model significantly outperformed adult-based echocardiography foundation models trained on substantially larger datasets (p < 0.001). Finally, our model demonstrated robust performance across patient age, patient sex, and studies with varying number of videos.

**Conclusions:** Our findings demonstrate the remarkable potential for multi-task deep learning models to aid the interpretation of pediatric echocardiograms. In addition, our results underscore the need for models that are specifically tailored to pediatric populations.

## 1 Introduction

Congenital heart defects afflict nearly 1% of all births worldwide and represent the most common category of birth anomaly [1]. While highly prevalent, these conditions comprise an exceptionally heterogeneous group of structural cardiac malformations that differ widely in anatomical complexity, physiological impact, and clinical course [2, 3]. Accurate diagnosis of congenital heart disease can be particularly challenging, as subtle structural abnormalities must be recognized within the rapidly changing context of pediatric cardiac growth and development. Subsequent surgical and catheter-based interventions further alter cardiac geometry and physiology, creating unique and highly complex postoperative anatomy which must be monitored over time [4, 5]. Echocardiography remains the principal modality for evaluation and follow-up due to its safety, accessibility, and ability to provide real-time assessment of cardiac structure and function [6, 7]. However, comprehensive interpretation of pediatric echocardiograms requires significant expertise and time, and the number of trained specialists is insufficient to meet clinical demand [8]. Moreover, substantial inter- and intra-observer variability persists even among experienced clinicians, contributing to variability in diagnostic accuracy and delays in patient evaluation [7, 9, 10].

Notably, deep learning (DL) models have demonstrated significant potential in automating and improving echocardiographic analyses [11, 12]. For adult populations, models have progressed from those trained on relatively small datasets restricted to a single task and specific echocardiographic views [13, 14] to large-scale systems capable of performing numerous tasks using a wider array of views [15–17]. However, similar progress has not been observed for pediatric populations. Pediatric echocardiography presents unique challenges due to a greater variation in heart rate, limited patient cooperation, and a wide spectrum of congenital and acquired heart diseases [18]. Consequently, models developed for adults have failed to generalize to pediatric populations [19]. Despite this, existing pediatric-based models remain limited to small training datasets, narrow diagnostic scopes, and specific echocardiographic views, thus limiting their broader clinical utility [19–21].

To address this gap, we introduce EchoAI-Peds: the first multi-task deep learning model for pediatric echocardiography. EchoAI-Peds is trained on the comprehensive set of pediatric labels to date and is designed to integrate information from multiple echocardiographic views simultaneously. Specifically, we develop a video-based vision transformer to simultaneously detect 28 congenital heart defects, structural and functional abnormalities, repairs, and interventions directly from complete pediatric echocardiography studies with multiple videos. To this end, we use over 700,000 pediatric echocardiogram videos from Stanford Medicine to develop EchoAI-Peds and test its efficacy on an internal, heldout dataset. In addition, we examine the generalizability of EchoAI-Peds through the analysis of a spatially and temporally distinct patient cohort at the Children’s Hospital of Philadelphia. To demonstrate the importance of pediatric-focused model development, we show that EchoAI-Peds outperforms existing foundation models for adult echocardiography that were trained on substantially larger datasets. Finally, we demonstrate that the performance of EchoAI-Peds is largely insensitive to the age and sex of the patient, as well as the number of videos within a given study.

## 2 Methods

### 2.1 Internal data source

We collected 14,552 pediatric echocardiography studies that were conducted between 2014 and 2021 at Stanford Medicine (SM) (IRB #52440), see **Figure 1A**. We included studies performed at the Lucile Packard Children’s Hospital as well as those from Stanford Healthcare in which patients were 18 years of age or younger. The resulting dataset included transthoracic, transesophageal, and stress echocardiograms, which contained standard 2D B-mode and color Doppler videos across a variety of echocardiographic views. In addition, every study was accompanied by a text report containing clinical findings verified by a board-certified cardiologist.

**Figure 1:**
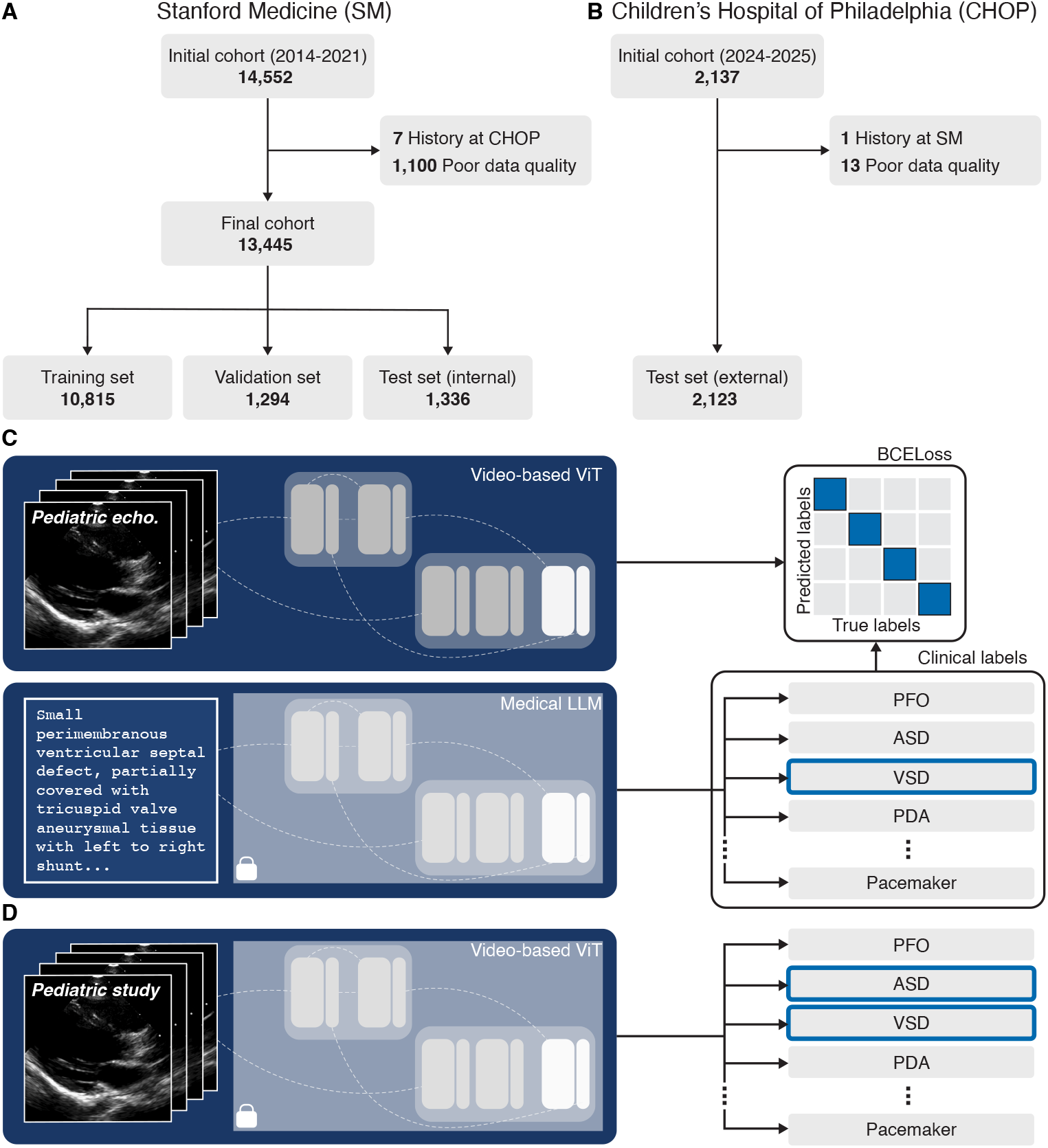
Overview of EchoAI-Peds framework. **a**. We collected pediatric echocardiography studies from Stanford Medicine (SM) for model development. We excluded any study from patients with a prior history at the Children’s Hospital of Philadelphia (CHOP) and those with inadequate echocardiogram videos. The remaining studies were split into training, validation, and internal testing sets. **b**. Similarly, we collected pediatric echocardiography studies at CHOP. We excluded any study with a prior history at SM as well as those with inadequate echocardiogram videos or reports. The remaining studies served as our external test set. **c**. For training, a single video was selected from each study. We trained the vision transformer (ViT) in EchoAI-Peds by minimizing the error between video-based label predictions and those extracted from clinical reports using a frozen, medically fine-tuned large language model (LLM). **d**. For inference, EchoAI-Peds integrated information from all available views within each study to simultaneously detect 28 clinical findings using the frozen, previously trained ViT.

### 2.2 External data source

At the Children’s Hospital of Philadelphia (CHOP), we collected 2,137 pediatric echocardiography studies that were conducted between 2024 and 2025 (IRB #25-023534), see **Figure 1B**. These included transthoracic echocardiograms, which contained standard 2D B-mode and color Doppler videos across a variety of echocardiographic views. Each study was accompanied by a text report containing clinical findings verified by a board-certified cardiologist.

### 2.3 Data pre-processing

To prevent potential data leakage between model training and external testing, we first removed studies of patients at SM with a clinical history at CHOP and vice versa. There were 7 such studies at SM and 1 such study at CHOP. Next, we removed studies that described more than one patient in the corresponding clinical reports. In addition, we removed videos that failed to process and those containing fewer than 16 frames to satisfy the input requirements of our deep learning model. Studies in which all videos met these exclusion criteria were removed in their entirety. These exclusions resulted in the removal of 1,100 studies from SM and 13 from CHOP. From each study, we then removed patient identifiers from both the videos and clinical text of each study through a rigorous de-identification procedure. Next, we used an open-source large language model (LLM) fine-tuned on medical data, MedGemma 27B [22], to extract clinical class labels from each clinical report. See Supplementary Table S1 for a summary of the LLM’s performance in extracting these labels. We used the identified labels for model development and evaluation. Finally, the SM dataset was split at the patient level into a training set of 6,549 patients (80%), a validation set of 819 patients (10%), and a held-out internal testing set of 819 patients (10%). This yielded 626,901 videos from 10,815 studies for training, 75,664 videos from 1,294 studies for validation, and 76,867 videos from 1,336 studies for internal testing. In contrast, we reserved studies from all 1,981 patients at CHOP for external testing of our model. To that end, we reserved 112,657 videos from 2,123 studies for this purpose. Further details on data pre-processing are found in Section 1.1 of the Supplementary Appendix.

### 2.4 Model training

We trained a video-based, multi-scale vision transformer [23] that simultaneously detects 28 clinical findings directly from pediatric echocardiogram videos, see **Figure 1C**. During training, we randomly selected a single video from each study, from which we then randomly sampled a 16-frame clip. This 16-frame clip was used as our model input to generate study-level predictions for all 28 clinical findings. Our model’s predictions were compared against the LLM-extracted labels to compute the training loss. We initialized our model with pretrained Kinetics weights [24] and optimized using the binary cross-entropy loss for 100 epochs with the AdamW optimizer and a learning rate of 1 × 10^−4^. Training was performed with an effective batch size of 64 on 2 NVIDIA H100 80GB graphics processing units. Following training, we selected the epoch checkpoint with the lowest validation loss for subsequent evaluation. Please refer to Section 1.2 of the Supplementary Appendix for more details on model development.

### 2.5 Model evaluation

We evaluated model performance on the SM and CHOP test sets using a multi-video, multi-clip strategy, see **Figure 1D**. Specifically, we randomly sampled up to four distinct 16-frame clips from each available video within a given study [17]. We first averaged predictions across all clips from the same video and subsequently averaged the predictions across all videos to generate the study-level predictions. Since the dataset was imbalanced across the 28 clinical classes, we evaluated model performance using the area under the receiver operating characteristic curve (AUROC). To benchmark our model’s performance, we compared our classification results against those of two state-of-the-art foundation models for adult echocardiography: EchoCLIP [15] and EchoPrime [16]. Please refer to Sections 1.3-1.5 in the Supplementary Appendix for more details on model evaluation, the video- and clip-based sensitivity analyses, and model sensitivity and specificity, respectively.

### 2.6 Subgroup analyses

Age and sex are known to significantly influence cardiac structure and function [25, 26]. Furthermore, different echocardiographic views capture distinct anatomical features of the heart [27, 28]. In addition, pediatric echocardiography studies significantly differ from those of adults [29]. Specifically, they may include a larger number of views due to the varied nature of congenital heart disease. In addition, they often contain sweeps centered around particular views to better visualize complex anatomy [29]. Moreover, the number and quality of acquired views vary across patients and sonographers [30]. In light of these variabilities, our model must also perform reliably across studies with differing numbers of views and videos. To assess this, we performed three distinct subgroup analyses based on patient age, patient sex, and the number of available echocardiogram videos per study for both the SM and CHOP test sets:

i. **Age-based Analysis:** We categorized studies into predefined groups according to the patient’s age (in years) at the time of examination (A): *A <* 1, 1 ≤ *A <* 3, 3 ≤ *A <* 8, 8 ≤ *A <* 12, 12 ≤ *A <* 18, and 18 ≤ *A*. [31].
ii. **Sex-based analysis:** We categorized studies by the patient’s recorded sex and excluded those without a recorded sex.
iii. **View-count-based analysis:** We categorized studies into predefined groups according to the number of available echocardiogram videos (N): *N <* 25, 25 ≤ *N <* 50, 50 ≤ *N <* 100, and 100 ≤ *N*.

In each of the above analyses, we included five clinical labels that had sufficient representation across all subgroups at both institutions, with a minimum of two positive cases per group. For each analysis, we computed AUROC values for our model.

### 2.7 Statistical analyses

We used nonparametric bootstrapping with 1,000 replicates to compute 95% confidence intervals for the AUROC values on the SM and CHOP test sets. Next, the same bootstrapping procedure was used to obtain two-sided p-values assessing whether the AUROC differences between EchoAI-Peds and each competing model (EchoCLIP and EchoPrime) were statistically significant. In addition, we applied the Bonferroni correction to control the family-wise error rate in performing multiple comparisons. Finally, this analysis was repeated for subgroups stratified by patient age, patient sex, and the number of available echocardiogram videos per study. The statistical analyses were conducted in R (version 4.5).

## 3 Results

### 3.1 Study population and model inference

EchoAI-Peds was developed and internally tested using data from Stanford Medicine. Specifically, our model was trained, validated, and internally tested on 626,901 videos from 10,815 studies, 75,664 videos from 1,294 studies, and 76,867 videos from 1,336 studies, respectively. These sets corresponded to 6,549 patients for training, 819 for validation, and 819 for internal testing. External testing was conducted on 112,657 videos from 2,123 studies corresponding to 1,981 patients at CHOP. Males are marginally overrepresented in the training (55%), validation (54%), and testing sets (53% at SM; 55% at CHOP). In the internal test set, the average patient age was 8.60 *±* 7.81 years, while in the external test set, it was 7.42 *±* 6.88 years. Patient demographics for all datasets are summarized in **Table 1**.

**Table 1:** EchoAI-Peds development and evaluation cohort characteristics. All data presented as n (%) or mean *±* SD.

| Characteristic | SM |  |  | CHOP |
| --- | --- | --- | --- | --- |
|  | Train | Val. | Test | Test |
| Patients | 6549 | 819 | 819 | 1981 |
| Studies | 10815 | 1294 | 1336 | 2123 |
| Age, years | 8.98 ± 7.81 | 8.92 ± 7.33 | 8.69 ± 7.34 | 7.42 ± 6.88 |
| *Sex* |  |  |  |  |
| Male | 3592 (55%) | 442 (54%) | 436 (53%) | 1098 (55.4%) |
| Female | 2957 (45%) | 377 (46%) | 383 (47%) | 881 (44.5%) |
| Unknown | — | — | — | 2 (0.1%) |
| *Congenital heart defects* |  |  |  |  |
| PFO | 1427 (13.2%) | 181 (14.0%) | 183 (13.7%) | 387 (18.2%) |
| ASD | 1159 (10.7%) | 117 (9.0%) | 171 (12.8%) | 154 (7.3%) |
| VSD | 1173 (10.8%) | 132 (10.2%) | 157 (11.8%) | 242 (11.4%) |
| PDA | 552 (5.1%) | 56 (4.3%) | 73 (5.5%) | 162 (7.6%) |
| BAV | 326 (3.0%) | 48 (3.7%) | 25 (1.9%) | 119 (5.6%) |
| ARD | 325 (3.0%) | 57 (4.4%) | 39 (2.9%) | 153 (7.2%) |
| TOF | 668 (6.2%) | 81 (6.3%) | 84 (6.3%) | 79 (3.7%) |
| PulmAtr. | 438 (4.0%) | 60 (4.6%) | 42 (3.1%) | 32 (1.5%) |
| DORV | 246 (2.3%) | 27 (2.1%) | 26 (1.9%) | 36 (1.7%) |
| TGA | 215 (2.0%) | 26 (2.0%) | 29 (2.2%) | 45 (2.1%) |
| MAPCAs | 308 (2.8%) | 45 (3.5%) | 34 (2.5%) | 19 (0.9%) |
| *Structural abnormalities* |  |  |  |  |
| LVDil. | 857 (7.9%) | 97 (7.5%) | 119 (8.9%) | 91 (4.3%) |
| LVH | 513 (4.7%) | 57 (4.4%) | 68 (5.1%) | 59 (2.8%) |
| RVDil. | 1111 (10.3%) | 117 (9.0%) | 141 (10.6%) | 257 (12.1%) |
| RVH | 984 (9.1%) | 111 (8.6%) | 132 (9.9%) | 204 (9.6%) |
| HCM | 254 (2.3%) | 24 (1.9%) | 45 (3.4%) | 19 (0.9%) |
| PE | 797 (7.4%) | 87 (6.7%) | 66 (4.9%) | 86 (4.1%) |
| *Functional abnormalities* |  |  |  |  |
| LVSysD. | 932 (8.6%) | 106 (8.2%) | 118 (8.8%) | 107 (5.0%) |
| LVDiaD. | 422 (3.9%) | 48 (3.7%) | 59 (4.4%) | 15 (0.7%) |
| RVSysD. | 671 (6.2%) | 62 (4.8%) | 60 (4.5%) | 125 (5.9%) |
| *Repairs and interventions* |  |  |  |  |
| ASD repair | 377 (3.5%) | 39 (3.0%) | 73 (5.5%) | 67 (3.2%) |
| VSD repair | 845 (7.8%) | 102 (7.9%) | 127 (9.5%) | 137 (6.5%) |
| PDA ligation | 302 (2.8%) | 35 (2.7%) | 29 (2.2%) | 40 (1.9%) |
| Glenn procedure | 431 (4.0%) | 45 (3.5%) | 49 (3.7%) | 89 (4.2%) |
| Fontan procedure | 292 (2.7%) | 34 (2.6%) | 30 (2.2%) | 76 (3.6%) |
| Heart transplant | 819 (7.6%) | 79 (6.1%) | 75 (5.6%) | 51 (2.4%) |
| Cardiac surgery | 3372 (31.2%) | 369 (28.5%) | 386 (28.9%) | 623 (29.3%) |
| Pacemaker | 338 (3.1%) | 10 (0.8%) | 27 (2.0%) | 22 (1.0%) |

EchoAI-Peds was trained to simultaneously detect 28 congenital heart defects, structural and functional abnormalities, repairs, and interventions directly from complete pediatric echocardiography studies with multiple videos. To obtain a study-level classification, we applied a multi-video, multi-clip strategy. Further details on model inference strategy are provided in Supplementary Section 1.3 and Supplementary Tables S2-S5.

### 3.2 Internal testing

On the internal test set, EchoAI-Peds achieved a macro-averaged AUROC of 0.91 (95% CI: 0.90-0.92). Our model accurately detected congenital heart defects, such as VSD with an AUROC of 0.89 (95% CI: 0.86–0.91), TOF with an AUROC of 0.97 (95% CI: 0.95–0.98), and TGA with an AUROC of 0.93 (95% CI: 0.90-0.95). It also performed strongly in detecting structural abnormalities, such as LVDil with an AUROC of 0.93 (95% CI: 0.90–0.95), RVDil with an AUROC of 0.92 (95% CI: 0.90-0.94), and HCM with an AUROC of 0.93 (95% CI: 0.87–0.98). In detecting functional abnormalities, our model also achieved high performance, including LVSysD with an AUROC of 0.94 (95% CI: 0.91-0.96), LVDiaD with an AUROC of 0.86 (95% CI: 0.82–0.91), and RVSysD with an AUROC of 0.94 (95% CI: 0.91-0.96). Finally, our model performed remarkably in detecting repairs and interventions, such as VSD repair with an AUROC of 0.95 (95% CI: 0.94–0.97), Glenn procedure with an AUROC of 0.99 (95% CI: 0.98-0.99), and heart transplant with an AUROC of 0.98 (95% CI: 0.96–1.00). See Supplementary Table S5 for a complete list of AUROC values with confidence intervals. In addition, see Supplementary Table S6 for a complete list of sensitivity and specificity values with confidence intervals.

### 3.3 External testing

On the external test set, EchoAI-Peds achieved a macro-averaged AUROC of 0.89 (95% CI: 0.88-0.90). Our model accurately detected congenital heart defects, such as VSD with an AUROC of 0.82 (95% CI: 0.80–0.85), TOF with an AUROC of 0.92 (95% CI: 0.90–0.94), and TGA with an AUROC of 0.88 (95% CI: 0.86-0.90). It also performed strongly in detecting structural abnormalities, such as LVDil with an AUROC of 0.91 (95% CI: 0.88–0.94), RVDil with an AUROC of 0.91 (95% CI: 0.89-0.92), and HCM with an AUROC of 0.95 (95% CI: 0.87–1.00). In detecting functional abnormalities, our model also achieved high performance, including LVSysD with an AUROC of 0.95 (95% CI: 0.93-0.97), LVDiaD with an AUROC of 0.81 (95% CI: 0.68–0.92), and RVSysD with an AUROC of 0.93 (95% CI: 0.91-0.95). Finally, our model performed remarkably in detecting repairs and interventions, such as VSD repair with an AUROC of 0.92 (95% CI: 0.89–0.95), Glenn procedure with an AUROC of 0.99 (95% CI: 0.99-0.99), and heart transplant with an AUROC of 0.99 (95% CI: 0.98–1.00). See Supplementary Table S7 for a complete list of AUROC values with confidence intervals.

Notably, model performance was comparable to that on the internal test set, see Figure 2. However, model performance in detecting MAPCAs was substantially lower on the external test set, with an AUROC of 0.73 (95% CI: 0.64–0.81) versus 0.92 (95% CI: 0.88–0.96) on the internal test set.

**Figure 2:**
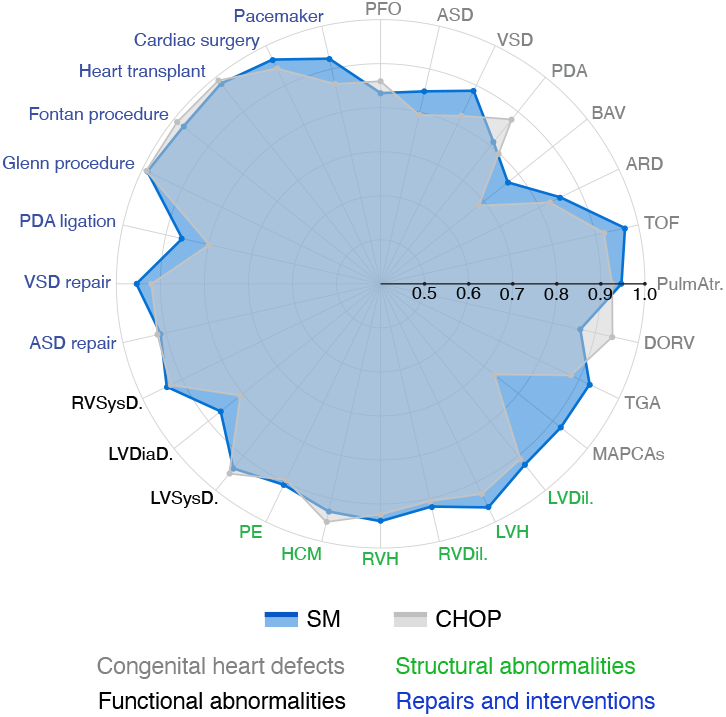
Comparison of EchoAI-Peds performance across institutions. Radar plot comparing AUROC values of EchoAI-Peds between the Stanford Medicine (SM) and the Children’s Hospital of Philadelphia (CHOP) test sets.

### 3.4 Comparison against adult echocardiography Models

We compared our model’s internal and external test performance to that of state-of-the-art foundation models for adult echocardiography: EchoCLIP [15] and EchoPrime [16]. On the internal test set, EchoCLIP and EchoPrime both obtained significantly lower macro-averaged AUROC values of 0.58 (95% CI: 0.56–0.60) and 0.58 (95% CI: 0.57–0.61), respectively (p *<* 0.001 for both). Notably, our model achieved higher performance against both models on all 28 clinical classes, see **Figure 3A**. Additionally, our model significantly outperformed EchoCLIP on all 28 clinical classes (p *<* 0.001 for all) and EchoPrime on 27 of the 28 clinical classes (p *<* 0.001 for 26 classes; p *<* 0.01 for 1).

**Figure 3:**
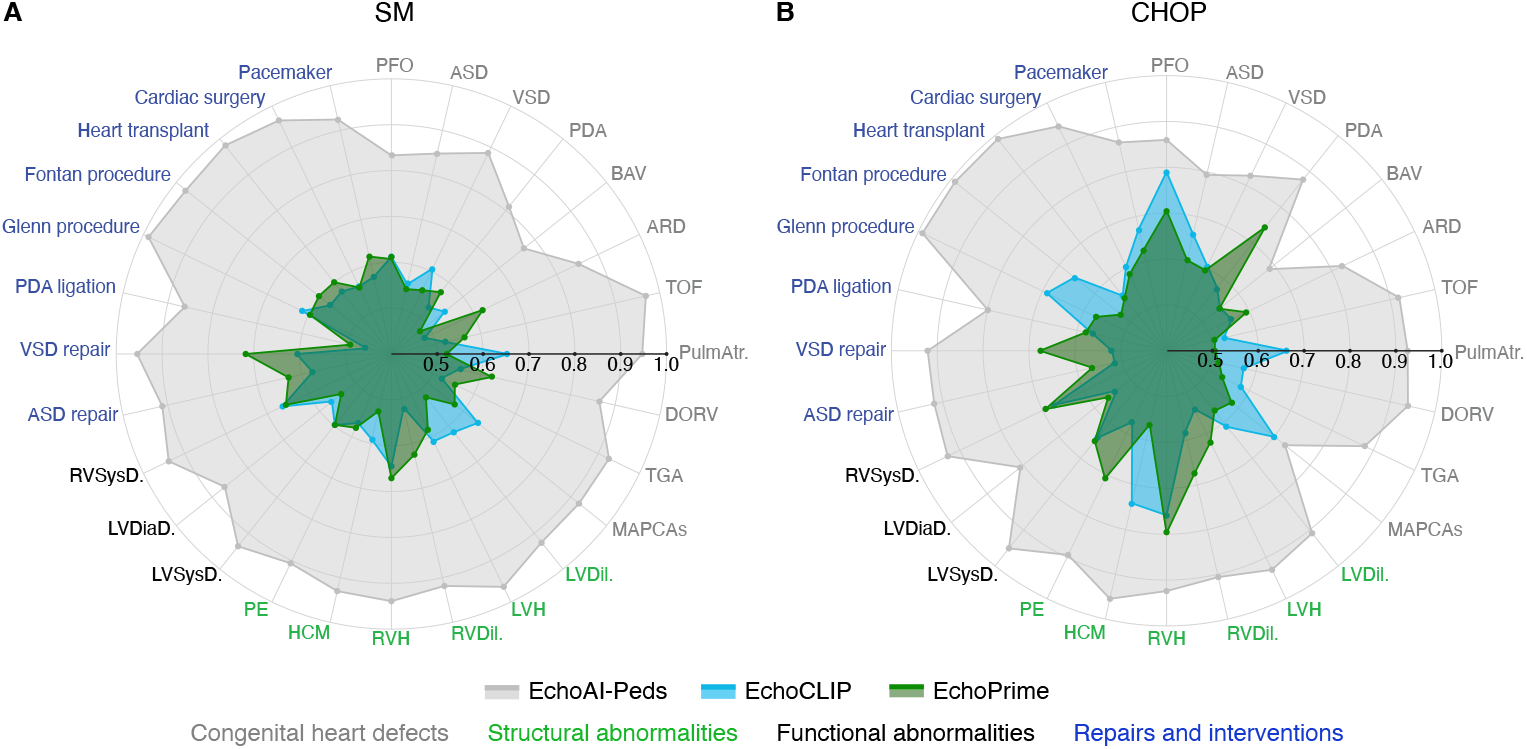
Comparison of EchoAI-Peds against adult-based models across institutions. Radar plots compare AUROC performance of EchoAI-Peds with foundation models for adult echocardiography (EchoCLIP and EchoPrime) on the **a**. Stanford Medicine (SM) test set and **b**. the Children’s Hospital of Philadelphia (CHOP) test set.

On the external test, EchoCLIP and EchoPrime also both obtained significantly lower macro-averaged AUROC values of 0.62 (95% CI: 0.60–0.63) and 0.61 (95% CI: 0.60–0.62), respectively. Notably, our model achieved higher performance against both models on all 28 clinical classes, see **Figure 3B**. Additionally, our model significantly outperformed EchoCLIP on 26 of the 28 clinical classes (p *<* 0.001 for 22 classes; p *<* 0.01 for three; p *<* 0.05 for 1) and EchoPrime on 26 of the 28 clinical classes (p *<* 0.001 for all 26). Complete lists of AUROC values with confidence intervals for model comparison on internal and external test sets are provided in Supplementary Tables S5 and S7, respectively.

### 3.5 Performance across subgroups

To test the sensitivity of our model to patient age, patient sex, and the number of available echocardiograms per study, we performed three distinct subgroup analyses for both the SM and CHOP test sets.

#### 3.5.1 Age-based results

On the internal test set, our model achieved consistent and strong results for VSD repair and cardiac surgery across all patient age groups. For VSD repair, AUROC values ranged from 0.93 for patients aged 12 to under 18 years to 0.96 for those younger than 1 year, aged 8 to under 12 years, and aged 18 years and older, see **Figure 4**A. In addition, for cardiac surgery, AUROC values ranged from 0.92 for patients younger than 1 year to 0.99 for those aged 18 years and older. Although our model performed strongly for LVDil, RVDil, and LVSysD across all age groups, its performance varied considerably by age. First, for LVDil, the AUROC value was 0.84 for patients younger than 1 year but 1.00 for those aged 18 years and older. Next, for RVDil, the AUROC value was 0.84 for patients younger than 1 year but 0.96 for those aged 12 to under 18 years. Finally, for LVSysD, the AUROC value was 0.84 for patients aged 18 years and older, but 0.98 for those aged 1 to under 3 years, aged 3 to under 8 years, and aged 8 to under 12 years.

**Figure 4:**
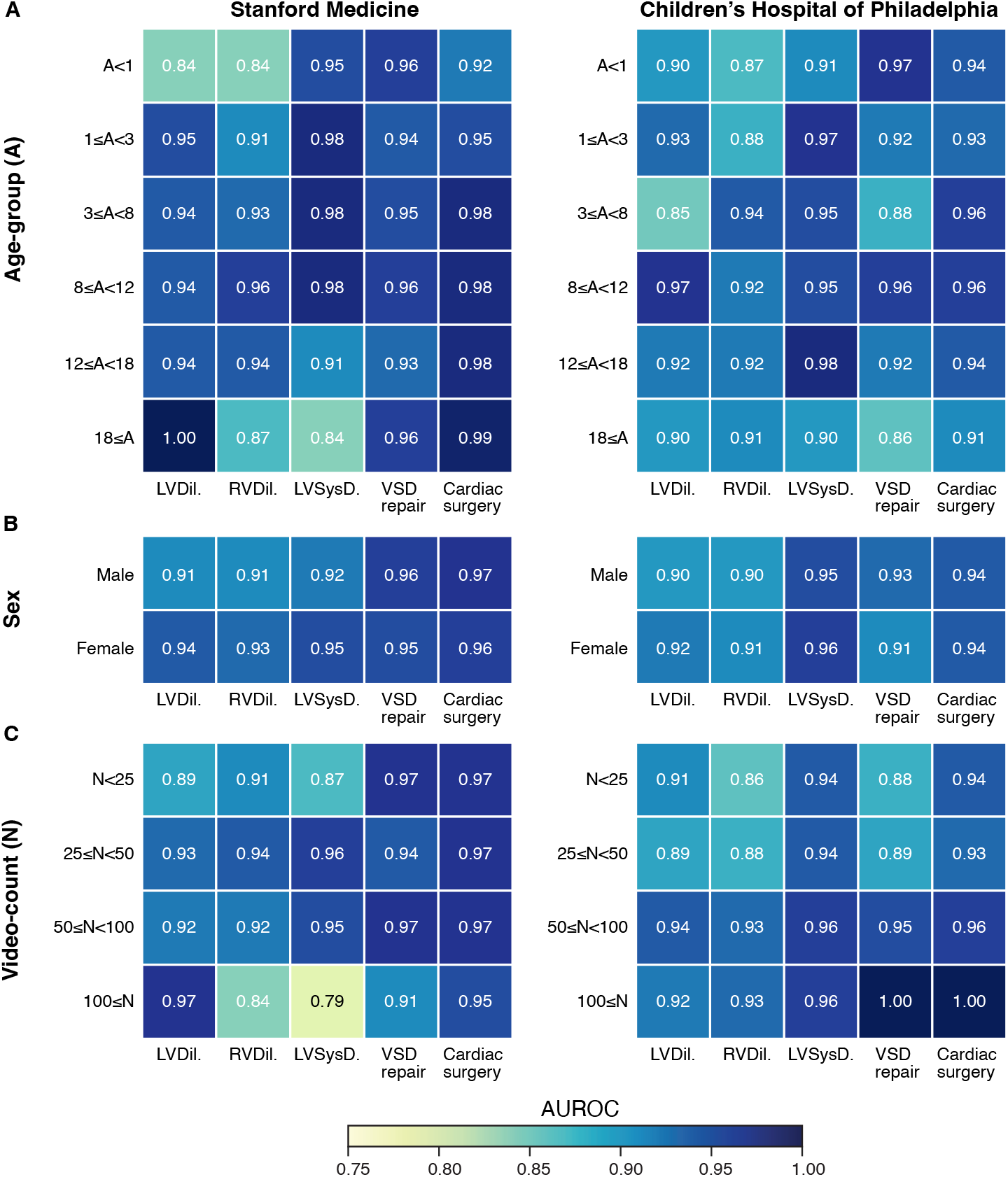
Comparison of EchoAI-Peds performance across subgroups. **a**. Square grid plots comparing AUROC values of EchoAI-Peds across distinct patient age groups. **b**. Square grid plots comparing AUROC values of EchoAI-Peds across patient sex. **c**. Square grid plots comparing AUROC values of EchoAI-Peds across studies with varying number of videos. The left and right plots show results on the Stanford Medicine (SM) test set and the Children’s Hospital of Philadelphia (CHOP) test set, respectively.

On the external test set, our model achieved consistently strong results for RVDil, LVSysD, and cardiac surgery across all age groups. For RVDil, the AUROC values ranged from 0.87 for patients younger than 1 year to 0.94 for those aged 3 to under 8 years. In addition, for LVSysD, AUROC values ranged from 0.90 for patients aged 18 years and older to 0.98 for those aged 12 years to under 18 years. Furthermore, for cardiac surgery, the AUROC values ranged from 0.91 for patients aged 18 years and older to 0.96 for those aged 3 to under 8 years and aged 8 to under 12 years. Although our model performed strongly for LVDil and VSD repair across all age groups, its performance varied considerably. First, for LVDil, the AUROC value was 0.85 for patients aged 3 to under 8 years but 0.97 for those aged 8 to under 12 years. Next, for VSD repair, the AUROC value was 0.86 for patients aged 18 years and older but 0.97 for those younger than 1 year. See Supplementary Table S8 for a complete list of AUROC values with confidence intervals.

Overall, our model demonstrated strong performance across patient age groups, suggesting that its performance was not dependent on age.

#### 3.5.2 Sex-based results

Despite being trained on fewer female patients, our model achieved slightly higher performance for female patients in 3 of the 5 clinical classes: LVDil, RVDil, and LVSysD, see **Figure 4B**. Notably, this result was observed across both the internal and external test sets. For example, on the internal test set, our model achieved an AUROC of 0.94 for females versus 0.91 for males in detecting LVDil. Similarly, on the external test set, our model achieved an AUROC value of 0.92 for females versus 0.90 for males in detecting LVDil. See Supplementary Table S9 for a complete list of AUROC values with confidence intervals. Overall, our model’s performance remained consistently strong across sex, with no evidence of decline in either group.

#### 3.5.3 Video-count-based results

We stratified our internal and external test sets into four groups: Group I – studies with fewer than 25 videos, Group II – studies with 25 or more videos but also fewer than 50 videos, Group III – studies with 50 or more videos but also fewer than 100 videos, and Group IV – studies with 100 or more videos. On the internal test set, our model achieved consistent and strong results for LVDil, VSD repair, and cardiac surgery, see **Figure 4C**. For LVDil, AUROC values ranged from 0.89 for Group I to to 0.97 for Group IV. In addition, AUROC values for VSD repair ranged from 0.91 for Group IV to 0.97 for Groups I and III. Furthermore, AUROC values for cardiac surgery ranged from 0.95 for Group IV to 0.97 for Groups I, II, and III. Although our model performed strongly for RVDil and LVSysD across most video-count groups, its performance varied considerably by group. First, the AUROC value for RVDil was 0.84 for Group IV, but 0.94 for Group II. In addition, the AUROC value for LVSysD was 0.79 for Group IV, but 0.96 for Group II.

On the external test set, our model achieved consistent and strong results for LVDil, RVDil, LVSysD, and cardiac surgery. For LVDil, AUROC values ranged from 0.89 for Group II to 0.94 for Group III. For RVDil, AUROC values ranged from 0.86 for Group I to 0.93 for Groups III and IV. For LVSysD, AUROC values ranged from 0.94 for Groups I and II to 0.96 for Groups III and IV. For cardiac surgery, AUROC values ranged from 0.93 for Group II to 1.00 for Group IV. However, our model demonstrated strong but inconsistent performance for VSD repair. Specifically, the AUROC value was 0.88 for Group I but 1.00 for Group IV. See Supplementary Table S10 for a complete list of AUROC values with confidence intervals.

Overall, our model achieved strong performance across video-count groups, suggesting that its performance was not dependent on the number of available videos within a given study.

## 4 Discussion

In this work, we introduce EchoAI-Peds, the first multi-task deep learning model for pediatric echocardiography. Specifically, we developed a video-based vision transformer capable of simultaneously detecting 28 congenital heart defects, structural and functional abnormalities, repairs, and interventions directly from complete pediatric echocardiography studies with multiple videos. During inference, EchoAI-Peds leverages information from multiple echocardiographic views simultaneously to produce unified, study-level predictions. To our knowledge, we curated the most comprehensive set of pediatric labels to date for deep learning model development and evaluation. As the number of clinical conditions increases, larger datasets are needed to provide sufficient samples for each condition. To address this, we also assembled the largest pediatric echocardiogram dataset to date, comprising the greatest number of video clips, studies, and patients. EchoAI-Peds was developed and internally tested using these data from Stanford Medicine. In addition, we assessed the generalizability of EchoAI-Peds on a spatially and temporally distinct patient cohort at the Children’s Hospital of Philadelphia. EchoAI-Peds achieved strong performance on both internal and external test sets, significantly outperforming adult-based echocardiography foundation models trained on substantially larger datasets. Finally, EchoAI-Peds demonstrated robust performance across patient age groups, patient sex, and varying numbers of videos per study.

Deep learning has made significant progress in medical image analysis for adult populations, ranging from zero-shot disease diagnosis [32] to accurate prediction of long-term time-to-event patient outcomes [33]. However, advances in pediatric populations remain severely limited. For example, Chen et al. developed a model capable of detecting congenital heart defects (CHDs) from pediatric electrocardiograms, but it could not differentiate between specific CHD types [34]. More recently, Mayourian and colleagues have used deep learning to identify CHDs from electrocardiograms, but have omitted pediatric repairs or interventions [35]. In pediatric echocardiography, Reddy et al. introduced a video-based model trained on approximately 6,000 videos to estimate left ventricular ejection fraction from apical four-chamber and parasternal short-axis views [19]. Similarly, Jiang et al. developed an image-based model trained on roughly 13,000 frames to detect congenital heart disease from one of seven standard views [36]. Notably, these models were limited to single tasks and specific echocardiographic views. Moreover, they were restricted to small dataset sizes in part due to their reliance on manually annotated labels from clinical experts, such as disease and view-based classifications and anatomical segmentation masks. In contrast, our work leverages recent advancements in open-source large language models [22, 37] to accurately extract a wide array of clinical class labels at scale. To our knowledge, we are the first to apply such an approach to pediatric echocardiography. This allowed us to use a larger dataset consisting of a wider array of clinical conditions to develop our multi-task model. Notably, many of the tasks our model performs, such as detecting MAPCAs or identifying a Fontan procedure, have not been previously attempted by any deep learning model and underscore our model’s clinical novelty. Moreover, EchoAI-Peds was trained on complete pediatric echocardiography studies encompassing a variety of views. As a result, our model is able to leverage all available videos within a given study to generate study-level predictions, similar to how clinical experts interpret an echocardiogram [38].

Similar to our work, Holste et al. developed a multi-task model for adult echocardiography [17]. However, their training labels were derived from structured echocardiography reporting fields that explicitly indicated the presence of specific conditions. This approach is not generalizable to hospitals or datasets that do not maintain structured echocardiographic data of this kind [39]. Additionally, Christensen et al. and Vukadinovic et al. developed two vision-language foundation models for adult echocardiography, EchoCLIP and EchoPrime, respectively. Although both models were trained on more than 200,000 echocardiography studies from approximately 100,000 patients, they performed poorly on our pediatric datasets. This demonstrates that adult-based models, despite being trained on immensely large datasets, cannot be directly applied to pediatric populations due to fundamental differences in cardiovascular features and clinical conditions. In our work, we highlight the importance of developing a comprehensive set of labels specific to pediatric populations and training models directly on pediatric data. Notably, our results demonstrate that achieving strong multi-task model performance on pediatric data does not require training datasets on the scale of hundreds of thousands of studies.

Our study is subject to several limitations. First, our model produces binary predictions even for conditions that are typically graded by severity, such as left ventricular dilation. Second, our model was trained on data from a single institution, which may limit its robustness across other pediatric populations. Third, although our model can detect numerous clinical findings, it currently omits several congenital and acquired heart diseases, as well as several repairs and interventions. Finally, our model cannot predict quantitative measurements from echocardiogram data, such as fractional shortening. Such measures are essential for a comprehensive echocardiographic assessment. In this work, we focused primarily on clinical conditions specific to pediatric patients, and therefore excluded measurement-based labels.

Future updates to EchoAI-Peds will include training on larger, multi-institutional datasets to improve generalizability and expand coverage across a wider spectrum of disease states. In addition, future work will focus on enabling our model to generate both categorical and quantitative predictions—such as disease severity and functional measurements—to further improve its clinical interpretability and utility.

In conclusion, our findings reveal that multi-task deep learning models hold significant potential for streamlining pediatric echocardiography analyses. Moreover, our results highlight the importance of developing echocardiography AI systems specifically tailored to pediatric populations. We hope this work will accelerate the development of pediatric-focused deep learning models with broad clinical utility.

## List of Abbreviations

SM: Stanford Medicine
CHOP: Children’s Hospital of Philadelphia
PFO: Patent foramen ovale
ASD: Atrial septal defect
VSD: Ventricular septal defect
PDA: Patent ductus arteriosus
BAV: Bicuspid aortic valve
ARD: Aortic root dilation
TOF: Tetralogy of Fallot
PulmAtr: Pulmonary atresia
DORV: Double outlet right ventricle
TGA: Transposition of the great arteries
MAPCAs: Major aortopulmonary collateral arteries
LVDil: Left ventricular dilation
LVH: Left ventricular hypertrophy
RVDil: Right ventricular dilation
RVH: Right ventricular hypertrophy
HCM: Hypertrophic cardiomyopathy
PE: Pleural effusion
LVSysD: Left ventricular systolic dysfunction
LVDiaD: Left ventricular diastolic dysfunction
RVSysD: Right ventricular systolic dysfunction
ASD repair: Atrial septal defect repair
VSD repair: Ventricular septal defect repair
PDA ligation: Patent ductus arteriosus ligation

## 5 Data availability and software

The pediatric echocardiogram studies from Stanford Medicine used to train, validate, and internally test our model are not publicly available due to patient privacy considerations. Similarly, the pediatric echocardiogram studies from the Children’s Hospital of Philadelphia used for external validation are not publicly available for the same reason. The inference code and model weights will be made publicly available upon publication.

## 6 Acknowledgments

We would like to thank the Stanford Sherlock cluster and the Respublica high-performance computing cluster at the Children’s Hospital of Philadelphia (CHOP) for providing computational resources and support that contributed to these research results. In addition, we would like to acknowledge Dr. Wensi Wu for her help in familiarizing our team with HPC resources at CHOP.

## 7 Footnote

Supplementary information will be made publicly available upon acceptance for final publication.

